# Therapeutic Anticoagulation Is Associated with Decreased Mortality in Mechanically Ventilated COVID-19 Patients

**DOI:** 10.1101/2020.05.30.20117929

**Authors:** Muoi A. Trinh, Daniel R. Chang, Usha S. Govindarajulu, Erica Kane, Valentin Fuster, Roopa Kohli-Seth, Sanam Ahmed, Matthew A Levin, Martin D. Chen

## Abstract

**Objective:** To evaluate differences in morbidity and mortality among mechanically ventilated patients with COVID-19 treated with therapeutic versus prophylactic anticoagulation.

**Methods:** We performed a retrospective review of 245 COVID-19 positive patients admitted to the ICU requiring mechanical ventilation from March 1, 2020 through April 11, 2020 at Mount Sinai Hospital. Patients either received therapeutic anticoagulation for a minimum of 5 days or prophylactic dose anticoagulation. Morbidity and mortality data were analyzed.

**Results:** Propensity score (PS) weighted Kaplan-Meier plot demonstrated a survival advantage (57% vs. 25%) at 35 days from admission to the ICU in patients who received therapeutic anticoagulation for a minimum of 5 days compared to those who received prophylactic anticoagulation during their hospital course. A multivariate Cox proportional hazard regression model with PS weights to adjust for baseline differences found a 79% reduction in death in patients who were therapeutically anticoagulated HR 0.209, [95% CI (0.10, 0.46), p < 0.001]. Bleeding complications were similar between both groups. A 26.7% [95% CI (1.16, 1.39), p< 0.001] excess mortality was found for each 1 mg/dL rise in serum creatinine over a 21-day period.

**Conclusions:** Therapeutic anticoagulation is associated with a survival advantage among patients with COVID-19 who require mechanical ventilation in the ICU.

## Introduction

Coronavirus (SARS-CoV-2) is a novel single-stranded RNA virus and the causative agent of the COVID-19 pandemic. COVID-19 is related to severe acute respiratory syndrome coronavirus-1 (SARS-CoV-1) and middle east respiratory syndrome coronavirus (MERS-CoV), which were responsible for epidemics of viral pneumonia in 2003 and 2012 respectively^1^. Like SARS-CoV-1 and MERS-CoV, the pathophysiology of severe disease caused by COVID-19 appears to be partly due to endothelial injury leading to severe inflammation and dysregulated thrombosis in both the lungs and extrapulmonary organs^1–14^.

In the lungs, the degree of hypoxemia is out of proportion to expected compliance changes and there is concomitant evidence of decreased hypoxic pulmonary vasoconstriction and increased alveolar dead space ventilation consistent with pulmonary vascular endothelial injury and thrombosis rather than acute respiratory distress syndrome (ARDS)^5–6^. This suggests that initiation of therapeutic anticoagulation (TA), rather than prophylactic anticoagulation (PA) may be necessary to prevent multi-organ failure and death, particularly for patients with severe disease. In light of these reports, as well as our own institutional experience with arterial, venous and intravascular device thrombosis, we developed a system wide protocol for therapeutic anticoagulation in critically ill COVID-19 patients with a low risk of bleeding complications. Some of our health system’s experience with this anticoagulation protocol was recently reported, demonstrating improvements in 21-day survival from day of hospital admission in 395 COVID-19 patients who were mechanically ventilated with anticoagulation^15^. We conducted a separate analysis of therapeutic anticoagulation for intubated patients in a single hospital within our health system. We hypothesized that therapeutic dose anticoagulation for at least five days would have a survival advantage over prophylactic dose anticoagulation, in mechanically ventilated patients using a risk-adjusted model. A minimum of five days therapy was chosen based on the clinical observation that for many patients it took between two to three days to achieve a therapeutic level of anti-coagulation based on coagulation laboratory results.

## Methods

Approval was obtained from Icahn School of Medicine at Mount Sinai Institutional Review Board (IRB-20–0342) to perform a retrospective review of all COVID-19 positive adult (age > 18) patients admitted to any of the ICU at The Mount Sinai Hospital between March 1, 2020 and April 11, 2020. All patients were confirmed to have COVID-19 by PCR from nasopharyngeal swab. Patients who died within five days of ICU admission were excluded. **(Appendix: Consort Diagram)**. Patients were eligible for treatment with therapeutic anticoagulation if they did not have any of the following contraindications: active bleeding or a platelet count < 50× 10^3^/L on ICU admission, a history of heparin induced thrombocytopenia, or a history of stroke within the past year.

Dosing of anticoagulation was based on ideal body weight and estimated glomerular filtration rate (eGFR). Therapeutic heparin dose involved infusions of 15u/kg/hr or greater with or without a heparin bolus of 80units/kg with the goal to achieve an activated prothrombin time of 70–100 seconds based on institutional protocol. Therapeutic enoxaparin dose was defined as 1mg/kg twice daily if the GFR was > 30ml/min or once daily if the GFR ≤ 30ml/min. Factor Xa levels were not routinely used to monitor the efficacy of anticoagulation with enoxaparin during the study. In the prophylaxis group, patients received heparin 5000 units subcutaneously two to three times daily, or enoxaparin 40mg twice daily if the GFR > 30ml/min or 40mg once daily if GFR ≤ 30ml/min. Newly initiated apixaban 2.5mg or 5mg twice daily was considered prophylactic dosing.

We reviewed the electronic medical record of each patient to collect data including demographics, comorbidities, laboratory values, and concurrent therapies (Table 1). Baseline laboratory data were abstracted from the day of or date closest to admission to the ICU. Baseline sequential organ failure assessment (SOFA) scores were calculated from the date of admission to the ICU. Concurrent medical therapies to treat COVID-19 based on hospital protocols were also documented. Mortality data was obtained by following patients for 35 days from ICU admission. Bleeding complications were defined by a physician or consultant evaluation for active bleeding and/or a decline in hematocrit requiring at least one unit of blood transfusion.

**Table 1:**
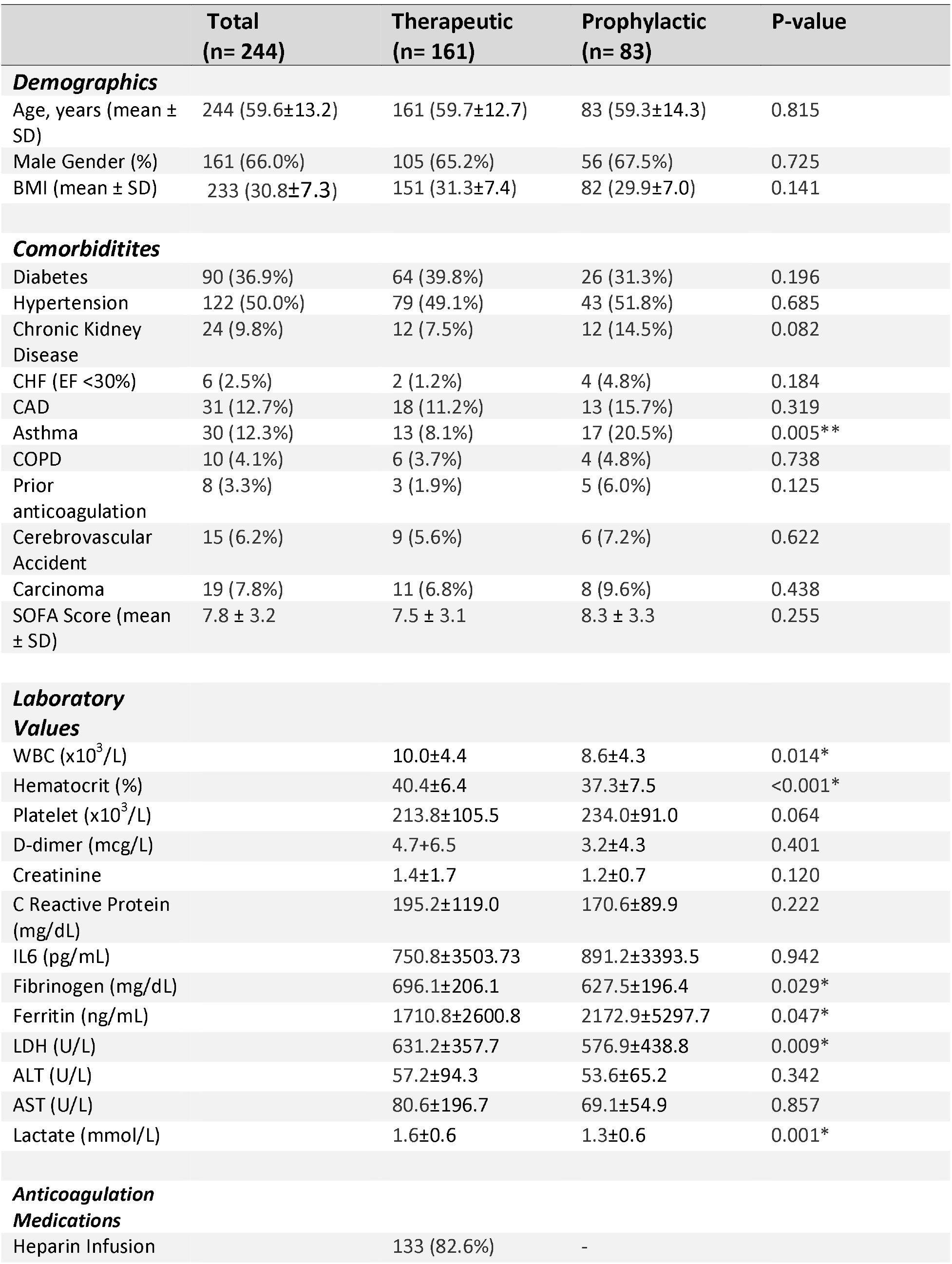
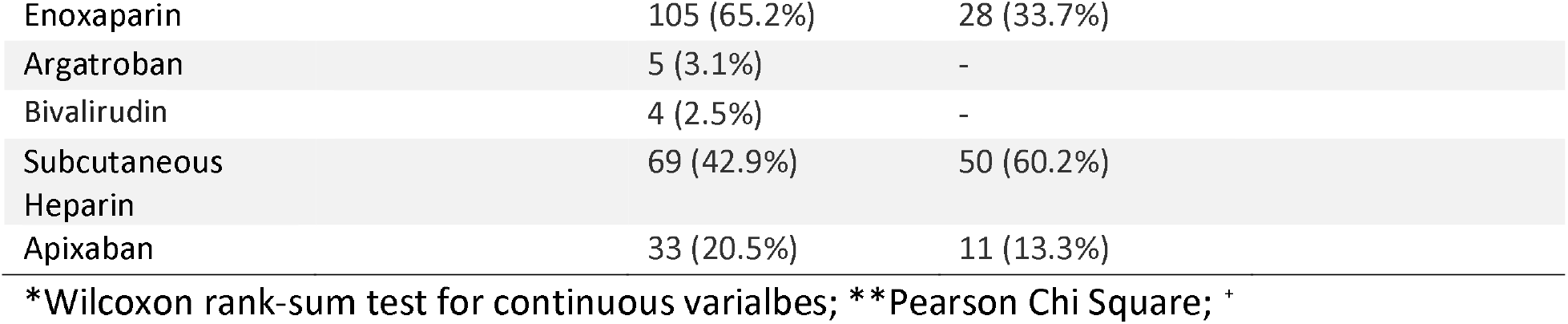
Baseline Characteristics

### Statistical Analysis

Univariate analysis was performed and reported as number and percentage for categorical variables. Mean and standard deviation or median and interquartile range were reported for continuous variables. The Mann-Whitney U test was used to compare groups for each continuous variable. Pearson’s chi-square or Fisher’s exact test were computed for contingency tables of each categorical variable by group. For survival analyses, time from ICU admission to date of death (or last date known alive) was used with right censoring and with left truncation for those who died less than five days after ICU admission. Kaplan-Meier (KM) plots were generated as unweighted, as well as, weighted by propensity scores (PS) generated from logistic regression with all covariates, to adjust for baseline differences between groups. For each patient, the weight was assigned as the inverse probability of belonging to the group that they are in (inverse probability of treatment weight). The KM curves were compared by the log-rank test. We ran a univariate and then followed with a multivariate Cox proportional hazards regression model with the PS weights to adjust for potential confounding variables in this time to death analysis and also to allow for creatinine to be a time-varying covariate in the model. All analysis was performed using SAS version 9.4 (SAS, Cary, NC). Statistical significance was set at p < 0.05.

## Results

During the study period, 279 patients were admitted to the ICU and required mechanical ventilation, of which 34 died within five days of admission. Thus 244 patients were included in the analysis: 161 received therapeutic anticoagulation and 83 received prophylactic anticoagulation. Baseline demographic and laboratory values are shown in **Table 1**. The majority of patients in the treatment group received a combination of therapeutic enoxaparin and heparin during their ICU course, rather than a single agent. On average, patients received therapeutic anticoagulation for seventeen days during their hospitalization. The patients in both groups had similar comorbidities including history of chronic kidney disease. Interestingly, patients in the therapeutic anticoagulation group had higher baseline lactate levels (1.6mmol/L) compared to the prophylaxis group (1.3mmol/L) (p< 0.001). However, Mann-Whitney U tests demonstrated that their baseline SOFA scores upon arrival to the ICU were comparable.

Patients with COVID-19 at our institution received multiple therapies based on protocols and these were documented for both groups **(Table 2)**. A slightly higher proportion of patients received corticosteroids 87.0% in the TA group compared to 75.9% in the PA group (p = 0.029). The PA group was twice as likely to receive tocilizumab as the TA group (p = 0.019). All other therapies were equally administered across the two groups.

**Table 2:** Concurrent Therapies

|  | **Total**<br>**n=244 (%)** | **Therapeutic**<br>**n=161(%)** | **Prophylactic**<br>**n= 83(%)** | **P-value** |
| --- | --- | --- | --- | --- |
| **Antimicrobials** | 242 (99.2) | 159 (98.8) | 83 (100.0) | 0.549 |
| **Corticosteroids** | 203 (83.2) | 140 (87.0) | 63 (75.9) | **0.029\*\*** |
| **Hydroxychloroquine** | 214 (88.4) | 137 (86.2) | 77 (92.8) | 0.127 |
| **Tocilizumab** | 35 (14.3) | 17 (10.6) | 18 (21.7) | **0.019\*\*** |
| **Sarilumab** | 21 (8.6) | 11 (6.8) | 10 (12.0) | 0.169 |
| **Remdesivir** | 11 (4.5) | 9 (5.6) | 2 (2.4) | 0.342 |
| **Serum Antibodies** | 8 (3.3) | 5 (3.1) | 3 (3.6) | 1.000 |
| **Stem Cell Therapy** | 10 (4.1) | 6 (3.7) | 4 (4.8) | 0.738 |
| **Anakinra** | 3 (1.2) | 3 (1.9) | 0 (0) | 0.553 |

\*\*Pearson Chi Square

Based on our survival KM analysis 58% of patients who received therapeutic anticoagulation survived to 35 days compared to 14% of patients in the prophylaxis group (p< 0.001). To adjust for baseline differences between the two groups a propensity score-weighted Kaplan Meier plot **(Figure 1)** was generated and showed a similar survival advantage for the anticoagulation treated group (57% vs. 25%, p < 0.001). Patients in the TA group had a longer length of stay with 18 days in the ICU compared to 11 days in the prophylactic group (Log rank test: p< 0.001), which may reflect increased survival.

**Figure 1:**
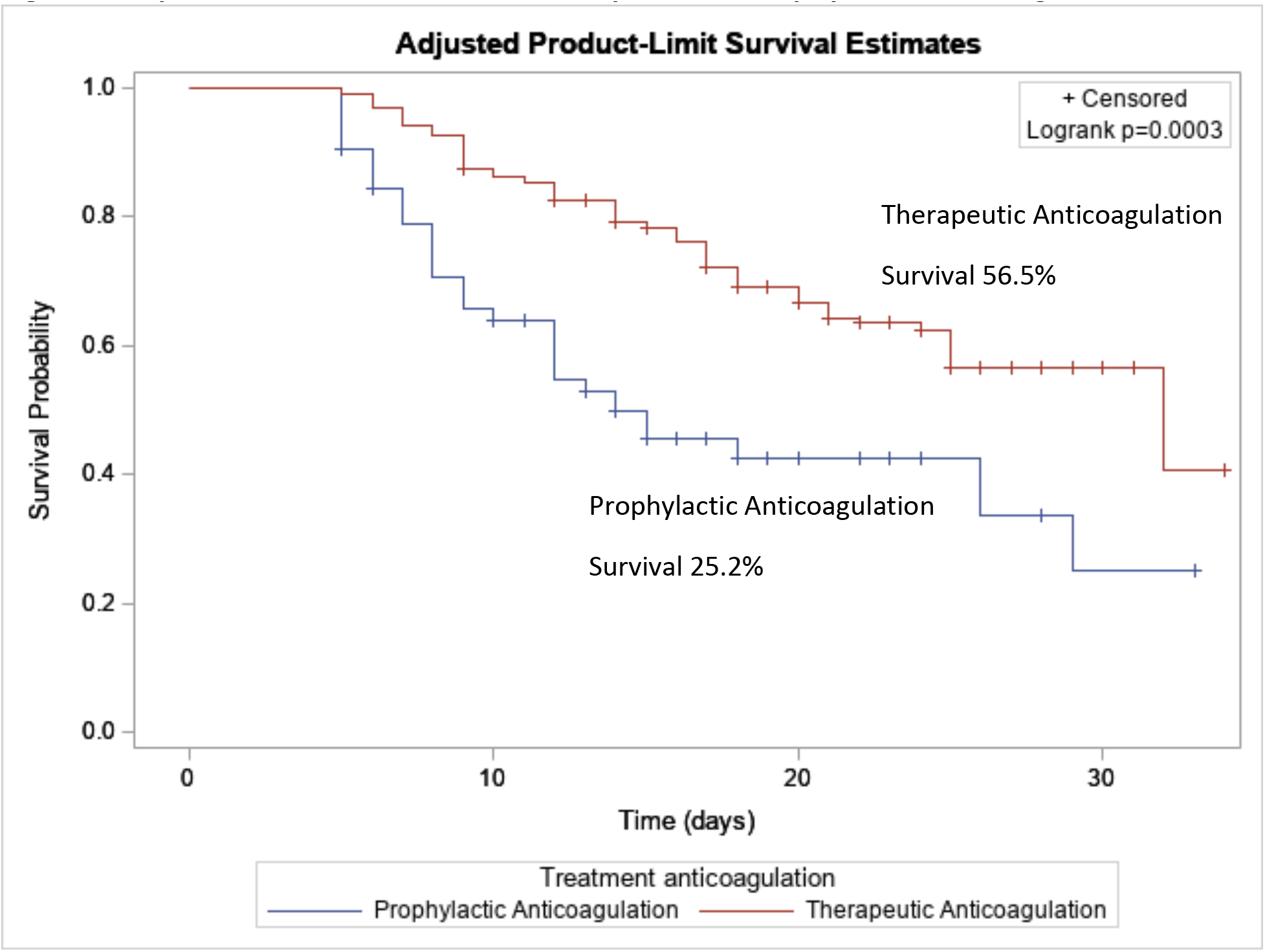
Kaplan-Meier Survival Curves, Therapeutic vs. Prophylactic Anti-coagulation. Patients were included for analysis in the treatment group if they received therapeutic doses of heparin or enoxaparin for a minimum of five days.

### Adverse Outcomes

There was no significant difference in adverse outcomes between the groups for stroke (3.7% vs 6.0%, p = 0.41) or liver failure (1.9% vs 2.4%, p = 1.0). There were trends towards increased risk of bleeding and renal failure requiring dialysis in the TA group, but neither reached statistical significance **(Table 3)**.

**Table 3:** Outcomes

|  | **Total**<br>**(n= 244)** | **Therapeutic**<br>**(n= 161)** | **Prophylactic**<br>**(n= 83)** | **P-value** |
| --- | --- | --- | --- | --- |
| **Stroke** | 11 (4.5%) | 6 (3.7%) | 5 (6.0%) | 0.41 |
| **Bleed** | 68 (27.9%) | 51 (31.7%) | 17 (20.5%) | 0.07 |
| **ESRD** | 92 (38.7%) | 67 (42.7%) | 25 (30.9%) | 0.08 |
| **Liver Failure** | 5 (2.1%) | 3 (1.9%) | 2 (2.4%) | 1.00 |
| **ICU LOS (mean days ± SD)** | 15.5±7.8 | 17.9±8.4 | 10.7±6.7 | **<0.001\*\*** |
| **Hosp LOS (mean days ± SD)** | 20.7±8.1 | 23.3±7.7 | 15.7±8.9 | **<0.001\*\*** |

\*Wilcoxon rank-sum test for continuous variables; \*\*Chi Square; Fischer's exact test

### Survival

A univariate Cox proportional hazards regression was performed with the group and the PS weights demonstrating treatment reduced mortality HR of 0.425 with a 95% C.I: (0.23, 0.78). We then ran a multivariate Cox proportional hazards regression model with the PS weights adjusted by these covariates: anticoagulation for 5 days, age, gender, history of chronic kidney disease, changes in creatinine over time, asthma, concurrent therapies (corticosteroids, tocilizumab), lactate, baseline SOFA score, and time from intubation day **(Table 4)**. We analyzed the effect of date of intubation, to account for differences in treatment of COVID-19 over time during the study period. In the model, therapeutic anticoagulation for at least five days reduced the rate of death by 79.1% [HR 0.209, 95% CI (0.10, 0.46), p < 0.001]. Older patients [HR 1.040, 95% CI (1.01, 1.07), p< 0.007] had a significantly increased rate of death.

**Table 4:** Propensity Score Weighted Cox Proportional Hazard Regression Model

|  | **Hazard Ratio** | **95%**<br>**Confidence Interval** | **P-value** |
| --- | --- | --- | --- |
| **Treatment anticoagulation**<br>**≥ 5 days** | 0.21 | 0.10-0.46 | **<0.001\*\*** |
| **Time Varying Creatinine** | 1.27 | 1.16-1.39 | **<0.001\*\*** |
| **Age** | 1.04 | 1.01-1.07 | **0.007\*\*** |
| **Gender (Female)** | 1.26 | 0.67-2.38 | 0.465 |
| **BMI** | 1.00 | 0.96-1.04 | 0.979 |
| **Asthma** | 0.67 | 0.20-2.20 | 0.515 |
| **Chronic Kidney Disease** | 0.46 | 0.16-1.35 | 0.160 |
| **Anticoagulation prior to**<br>**admit** | 0.19 | 0.01-5.60 | 0.337 |
| **Days to Intubation** | 1.00 | 0.95-1.06 | 0.933 |
| **Lactate** | 1.22 | 0.77-1.93 | 0.408 |
| **SOFA Score** | 1.08 | 0.97-1.20 | 0.141 |
| **Corticosteroids** | 1.86 | 0.70-4.97 | 0.213 |
| **Tocilizumab** | 1.44 | 0.59-3.47 | 0.422 |

\*\*Wald Chi Square

The absolute rate of death in the TA group was lower than the PA group, 34.2% vs. 53.0% (p< 0.005). Lastly, in terms of the changes in creatinine over a 21-day period after admission to the ICU, we found that mortality increased by 26.7% (p<0.001) for each g/dL unit increase. All other covariates including gender, prior anticoagulation, chronic renal disease, corticosteroid use, date of intubation, and laboratory values did not have statistically significant hazard ratios in the model. For the analyses, we assumed all available cases due to the pattern of missingness not being at random in most of the lab measures, which violated using an imputation procedure.

## Discussion

In this retrospective cohort study, we found that therapeutic anticoagulation was associated with markedly reduced mortality in intubated patients with COVID-19. This effect persisted when adjusted for confounding using propensity weights. Although we observed a slight trend towards increased bleeding complications in the TA group, this effect was not statistically significant. These findings suggest that the benefits of initiating therapeutic anticoagulation in intubated patients with COVID 19 outweighs the risk of bleeding.

There is abundant evidence supporting the assertion that severe COVID-19 involves dysregulated inflammation leading to pulmonary and extrapulmonary vascular endothelial injury and thrombosis^2,11,16–19^. While there is active research evaluating thrombolysis as an intervention for respiratory failure in COVID-19^20,21^, the effect of thrombolysis does not appear to be durable^20^. To date, there has been little conclusive evidence to date that the benefits of therapeutic anticoagulation outweigh the risks in COVID patients^19^. One previous study evaluated the benefit of anticoagulation in critically ill patients and found a benefit among a subset of critically ill patients with elevated sepsis-induced coagulopathy (SIC) scores and D- dimers, however this report used mostly prophylactic doses of anticoagulants and did not stratify for critically ill patients^22^. More recently, Paranjpe et al, analyzed outcomes for 2,773 COVID-19 patients treated throughout our hospital system and evaluated the effect of anticoagulation on mortality^15^. This group reported that therapeutic anticoagulation was associated with increased rates of mechanical ventilation, which is likely the result of a systemwide anticoagulation protocol in which all patients admitted to the ICU were placed on therapeutic anticoagulants. Our analysis confirms these findings while offering a greater level of detail on the exact nature of anti-coagulation used and the adverse outcomes.

### Limitations

There are several limitations to this study. First, due to its observational nature, there exists the potential for confounding. Although we attempted to control for this using a propensity matched model, the study took place during the COVID epidemic in NYC, when adjustments to our protocols and guidelines for therapy were constantly made as new information became available. The introduction of our anticoagulation protocol was one of these changes. We were concerned that the improved outcomes in the TA group could represent improvements in therapy as we gained experience managing these patients. We attempted to examine the effect of timing by analyzing the effect of date of intubation on outcome, which was not significant. Another limitation is our decision to exclude patients who died within 5 days of ICU admission (∼10% of ICU admissions). This potentially introduces immortal time bias.^23,24^ We felt however that including these patients would have introduced its own bias, since many of these patients died before having a chance to achieve full anti-coagulation, by falsely elevating the number of deaths among patients who were anticoagulated

## Conclusion

We report a significant decrease in mortality among these patients when treated with five days of therapeutic anticoagulation after intubation and demonstrate that survival advantage persisted when adjusted for potential confounders such as baseline covariates and concurrent therapies. We believe that these findings suggest that the initiation of therapeutic anticoagulation for critically ill patients with COVID-19 who require mechanical ventilation is a beneficial intervention.

## Data Availability

A de-identified data set may be made available upon request, for reproducibility or collaborative purposes only, subject to approval by the Mount Sinai Data Use Committee.

## Appendix

**Appendix 1:**
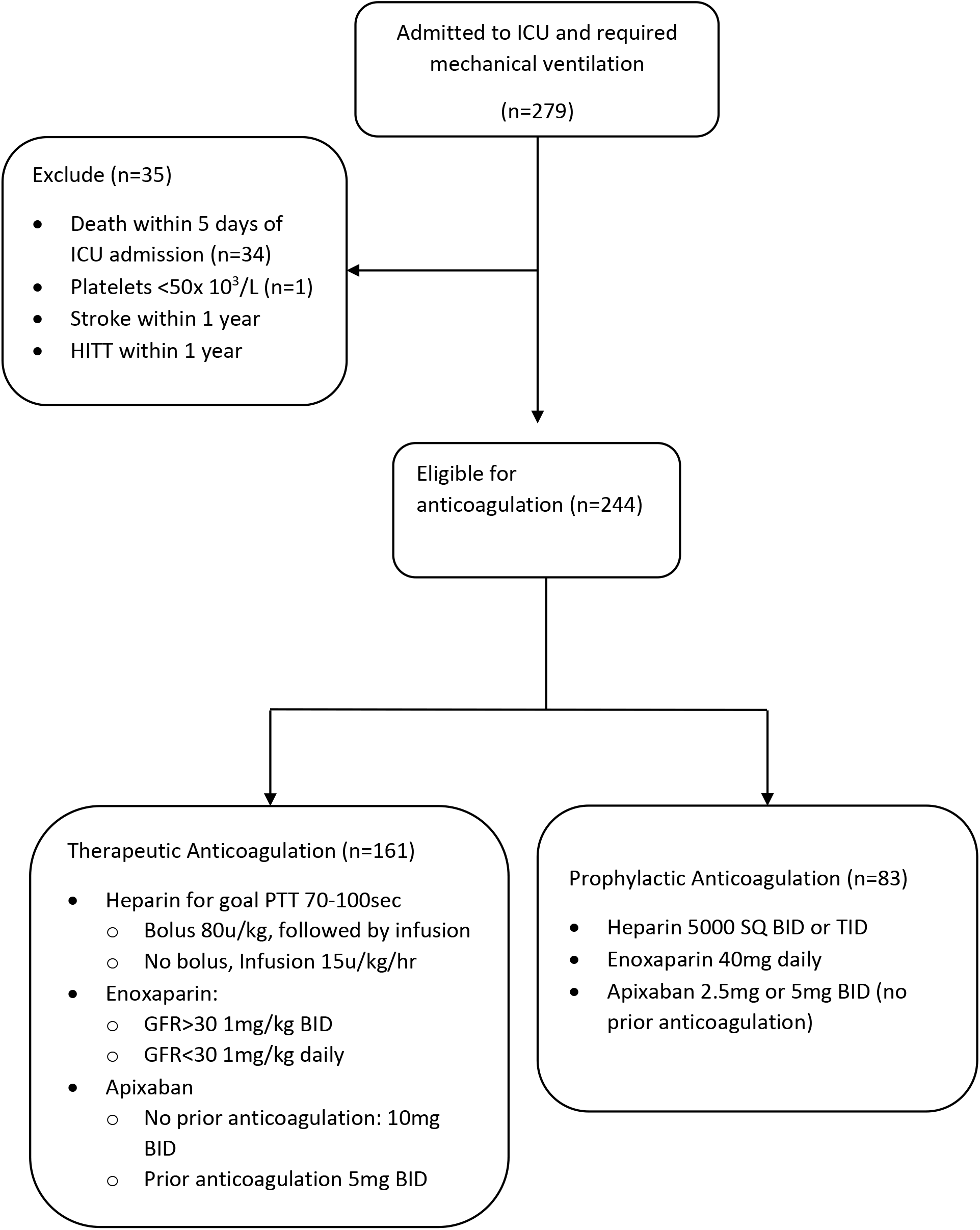
Consort Diagram

## Notes

### Competing Interest Statement

The authors have declared no competing interest.

### Clinical Trial

Retrospective Observational Study

### Funding Statement

Internal/Departmental Funding

### Author Declarations

Program for the Protection of Human Subjects, Icahn School of Medicine at Mount Sinai, New York, NY 10029

